# Novel *SYK* variant causes enhanced SYK autophosphorylation and PI3K activation in an antibody-deficient patient

**DOI:** 10.1101/2025.04.10.25325434

**Authors:** Emily S.J. Edwards, Josh Chatelier, Gregory I. Snell, Go Hun Seo, Rin Khang, Robyn E. O’Hehir, Julian J. Bosco, Menno C. van Zelm

## Abstract

**Background:** Inborn errors of immunity (IEI) affecting B-cell receptor signaling cause predominantly antibody deficiency (PAD) with varying degrees of severity. Recently, four heterozygous variants in *SYK* were reported to cause hypogammaglobulinemia, multiorgan inflammatory disease and diffuse large B-cell lymphoma.

**Objective:** We aimed to unravel the genetic and functional cause of PAD in a female (40-45 years) presenting with hypogammaglobulinemia, congenital heart disease and pulmonary hypertension requiring lung transplantation.

**Methods:** Patient gDNA was subjected to whole-exome and Sanger sequencing. Blood B- and T-cell subsets, as well as tonic and antigen-receptor induced expression levels of phosphorylated-SYK, phosphorylated-ribosomal S6 and phosphorylated p38 were evaluated by flow cytometry.

**Results:** A novel heterozygous missense *SYK* variant was identified, mutating a residue in the protein kinase domain (c.1769G>A; p.R590Q), which is highly conserved across vertebrates. While total B- and T-cell numbers were within the normal range, the patient had reduced unswitched and class-switched memory B-cell numbers. Resting B cells from the patient demonstrated enhanced autophosphorylation of SYK, and tonic and ligand-induced phospho-S6 levels. Spontaneous SYK autophosphorylation, S6 and p38 phosphorylation were recapitulated in a pre-clinical cell model, i.e. expression of the SYK R590Q variant in HEK293T cells.

**Conclusions:** We identified a novel gain-of-function variant in *SYK* to underlie hypogammaglobulinemia and atypical autoinflammatory disease. Flowcytometric screening for phospho-S6 in lymphocytes of IEI patients can guide genetic diagnosis of B-cell signaling abnormalities.

**KEY MESSAGES:**

- A novel monoallelic missense variant in *SYK* is identified in a patient with hypogammaglobulinemia and atypical autoinflammatory disease.
- Increased SYK autophosphorylation and enhanced tonic PI3K and MAPK signaling are indicative of a gain-of-function effect.
- Flowcytometric detection of phosphorylated S6 can provide a rapid functional evaluation of genetic variants affecting B-cell receptor signaling.

**Capsule summary:** Novel *SYK* variant enhances tonic PI3K signaling in human B cells, resulting in hypogammaglobulinemia.

## INTRODUCTION

Spleen tyrosine kinase (SYK) is a 72kDa non-receptor tyrosine kinase that critically mediates signaling downstream of immune receptors bearing immunoreceptor tyrosine-based activation motifs (ITAMs), such as the B-cell antigen receptor (BCR) and Fc receptors.[1, 2] Upon receptor engagement, tyrosine phosphorylation of ITAMs occurs, with the ITAM serving as a docking site for SYK SH2 domains. The docking relieves SYK autoinhibition and at least 10 different tyrosine residues are phosphorylated, including Tyr-323, Tyr-352 and Tyr-525/526.[2, 3] SYK autophosphorylation results in direct binding of downstream effector proteins, such as the p85α subunit of phosphoinositol-3-kinase (PI3K), Subsequent p85α phosphorylation by SYK activates the PI3K pathway, which is crucial for immune cell development, differentiation and function.[4–6] SYK is predominantly expressed in B cells, macrophages and plasma cells, with low levels found in mature CD4^+^ and CD8^+^ T cells.[7, 8] Additionally, SYK expression in endothelial cells mediates pulmonary vasoconstriction and vasodilation, and in osteoclasts it mediates bone resorption.[9–11]

Recently, four unique monoallelic gain-of-function *SYK* variants were identified in six patients with hypogammaglobulinemia, multiorgan inflammatory disease and diffuse large B-cell lymphoma.[12] Here, we describe a novel pathogenic, monoallelic variant in *SYK* in an adult presenting with hypogammaglobulinemia and atypical immune dysregulation.

## RESULTS AND DISCUSSION

Our case is a female in her fourties with delayed diagnosis of an inborn error of immunity (IEI). She initially presented with an incidental finding of hypogammaglobulinemia at 40-45 years of age during assessment for lung transplantation (LTx) in the setting of severe pulmonary arterial hypertension, accredited at the time to the delayed repair to a patent ductus arteriosis (**Table 1 and Supp Table E1**). The hypogammaglobulinemia prompted referral to a clinical immunologist, who noted a minimal prior infection history with genital herpes and infrequent episodes of recurrent sinusitis and otitis in adulthood. Examination did reveal digital clubbing, baseline hypoxia requiring oxygen supplementation and low set ears. Serological analysis demonstrated low serum IgG levels (**Supplementary Table E1**), undetectable serology for hepatitis B, C, HIV and CMV, and a poor response to pneumococcal vaccination (full Clinical history in the article’s Online Repository).

**Table 1.** Clinical characteristics of SYK GOF patients. CNS, central nervous system; DLBCL, Diffuse large B-cell lymphoma

|  | **This report**<br>**(n=1)** | **Wang *et al.*, 2021**<br>**(n=6)** |
| --- | --- | --- |
| Age of first presentation | 40-45y | 2wk – 44yr |
| Sex | F | F (66%) |
| Consanguineous | No | No |
| Parental evidence of immunodeficiency | No | No/unknown |
| Hypogammaglobulinemia | + | 6/6 (100%) |
| Recurrent Infections | - | 6/6 (100%) |
| Inflammation of: |  |  |
| Intestines | + | 6/6 (100%) |
| Skin | - | 6/6 (100%) |
| Joints | - | 4/6 (66%) |
| Lung | + | 3/6 (50%) |
| CNS | - | 2/6 (33%) |
| Liver | - | 1/6 (17%) |
| DLBCL | - | 2/6 (33%) |
| Pulmonary hypertension | + | - |
| SYK variant(s) | c.1769G>A<br>(p.R590Q) | c.1024C>A, c.1057G>A,<br>c.1350G>A, c.1649C>A,<br>c.1649C>T<br>(p.P342T, p.A353Y, p.M450I,<br>p.S550Y, p.S550F) |
| Autophosphorylation of SYK | Increased | Increased (6/6, 100%) |
| Vaccination response | Poor pneumococcal response | Not defined |
| Alive at time of report | Yes | Yes (83%) |

CNS, central nervous system; DLBCL, Diffuse large B-cell lymphoma

The patient received a bilateral lung transplant one year after diagnosis of hypogammaglobulinemia. The perioperative course was complicated by hemorrhage, pulmonary emboli and acute renal failure. In the absence of clinical infection, IgG replacement was delayed until post-LTx, and initiated alongside standard triple immunosuppression. The native lung explant noted severe pulmonary hypertension changes with additional thromboembolism and unusual diffuse alveolar damage with hemorrhage. Following LTx, the patient returned to full health, although she presented with premature ovarian insufficiency, a colonic ulcer and pancreatic pseudocysts. Three years post-LTx, she suffered from a severe picornavirus pneumonia, followed by progressive diffuse fibrotic lung disease and respiratory failure. The patient was re-transplanted with a single right lung. Currently ∼8 months later, the patient is doing well (**Table 1 and Supp Table E1**).

The hypogammaglobulinemia, autoinflammation and lung complications post-transplant prompted further evaluation of an underlying immune disorder, which included whole-exome sequencing (WES). WES yielded 24,798 homozygous and 41,989 heterozygous variants. Of these, 54 homozygous and 2967 heterozygous variants were deemed to be rare (allele frequency ≤0.01 in gnomAD), and of these, 68 deemed as loss-of-function and 605 as missense (**Figure 1A**). The missense variants included a novel heterozygous variant in exon 12 of *SYK* (c.1769G>A, **Figure 1B,C**), which was absent from gnomAD and predicted to be damaging by CADD (25.8), SIFT (deleterious) and PolyPhen (probably damaging). Presence of this variant in the patient was confirmed by Sanger Sequencing (**Figure 1B**), while the variant was not detected in her unaffected sibling or a healthy control (**Figure 1B**). Both parents did not consent to clinical, immunological or genetic evaluation. The variant affected a highly conserved residue within the SYK protein kinase domain (**Figure 1C,D**).[13]

**Figure 1:**
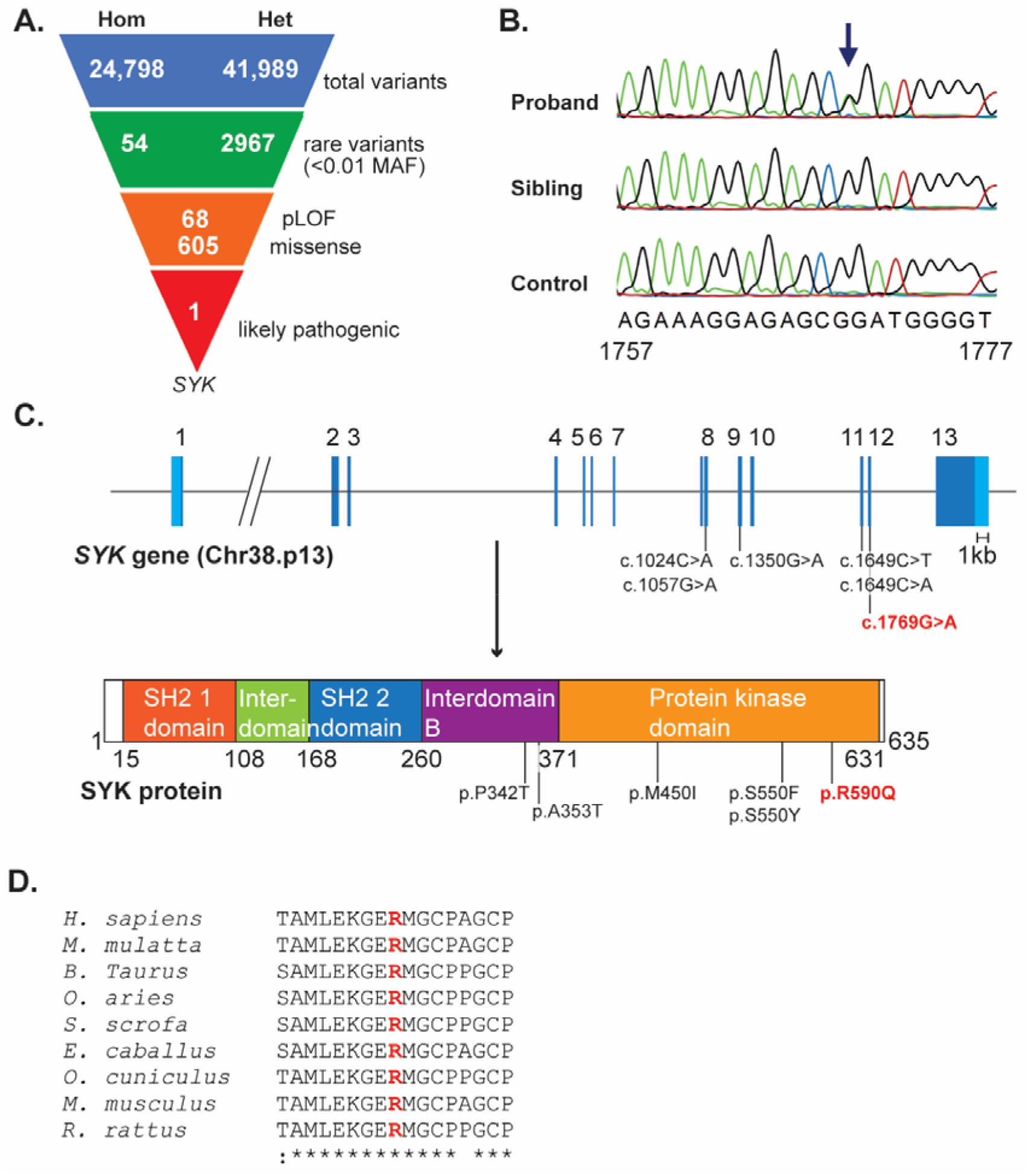
Identification of heterozygous missense variant in *SYK* in the patient. (**A**) Filtering of variants identified by whole-exome sequencing. (**B**) Sanger sequencing confirmation in patient, unaffected sibling and healthy control. (**C**) Schematics of the *SYK* gene and SYK protein annotated with variant identified in our patient and the 5 previously reported in SYK GOF patients.[12] (**D**) Multispecies alignment of the SYK protein kinase domain which includes the R590 residue.

The patient’s immune cell numbers pre-LTx were within the normal range, except for reduced IgG^+^CD27^+^ memory B cells. 31 months after receiving the LTx, the patient had increased granulocytes and dramatically reduced lymphocyte numbers, likely a result of the immunosuppression regimen to prevent organ rejection. Her TCRγ*δ*^**+**^ T cells, total CD4^+^ and CD8^+^ T cells and B cells were reduced, with memory B-cell numbers most dramatically affected (**Table 2**).

**Table 2.**
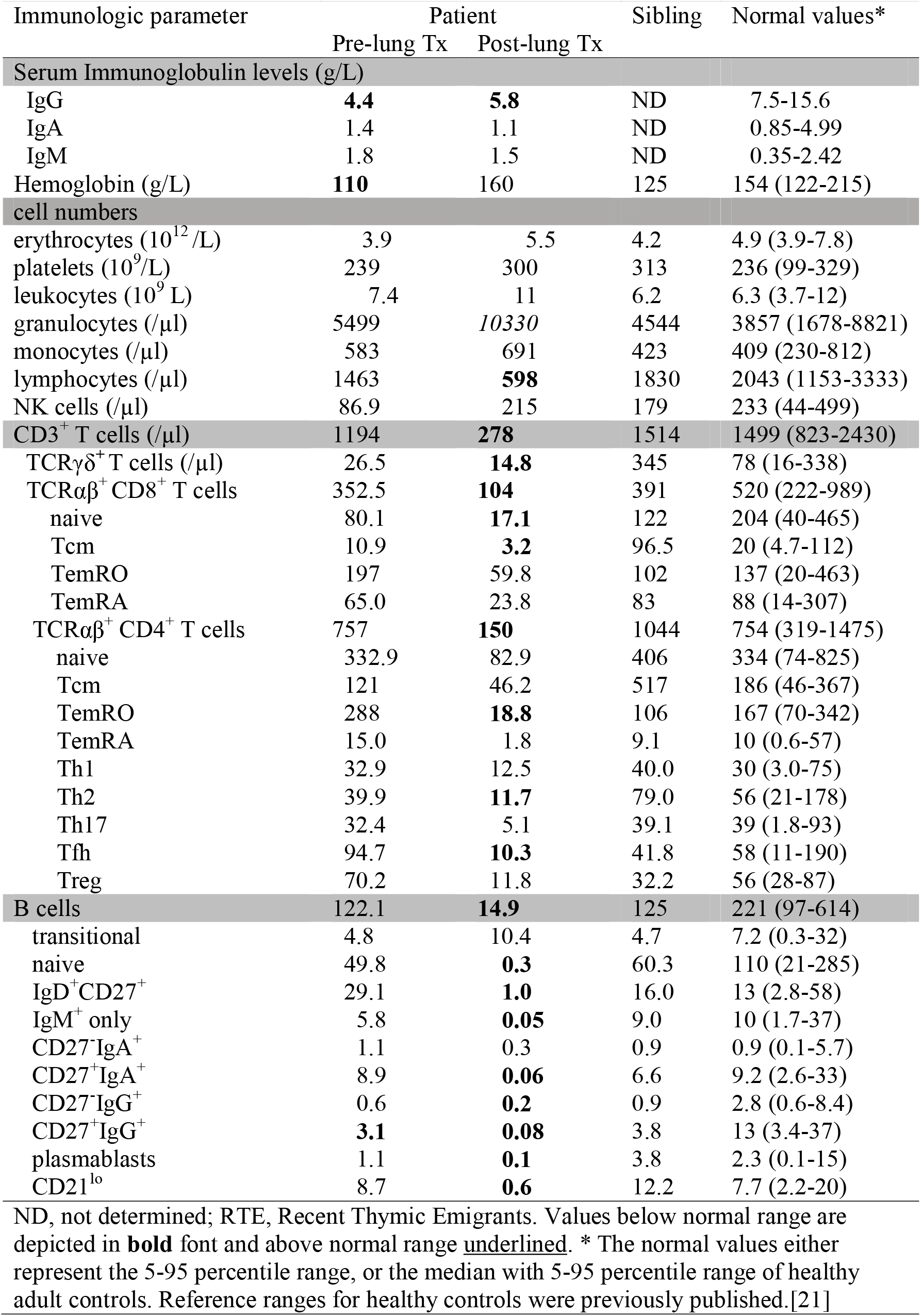
Immunological findings of SYK GOF patient. Immunologic parameter Patient Sibling Normal values* Pre-lung Tx Post-lung Tx ND, not determined; RTE, Recent Thymic Emigrants. Values below normal range are depicted in **bold** font and above normal range <u>underlined</u>. * The normal values either represent the 5-95 percentile range, or the median with 5-95 percentile range of healthy adult controls. Reference ranges for healthy controls were previously published.[21]

As the B-cell antigen receptor (BCR) signals through SYK to engage the PI3K pathway (**Figure 2A**), the effects of the p.R590Q variant were evaluated in stored B cells from a pre-LTx sample of the patient with and without anti-IgM stimulation (**Figure 2B-G**). In the absence of BCR stimulation, the patient’s B cells showed ∼8-fold higher autophosphorylation of SYK (phospho-SYK) than the healthy control and the unaffected sibling (**Figure 2B**). Upon anti-IgM stimulation, similarly high levels of phospho-SYK were detected in B cells of the patient, healthy control and the unaffected sibling (**Figure 2C**). Resting and CD3-stimulated T cells from all individuals had undetectable levels of phospho-SYK (**Figure 2B&C**), in line with the low expression of SYK in mature T cells.[14] Thus, the p.R590Q variant is associated with enhanced autophosphorylation of SYK in resting B cells.

**Figure 2:**
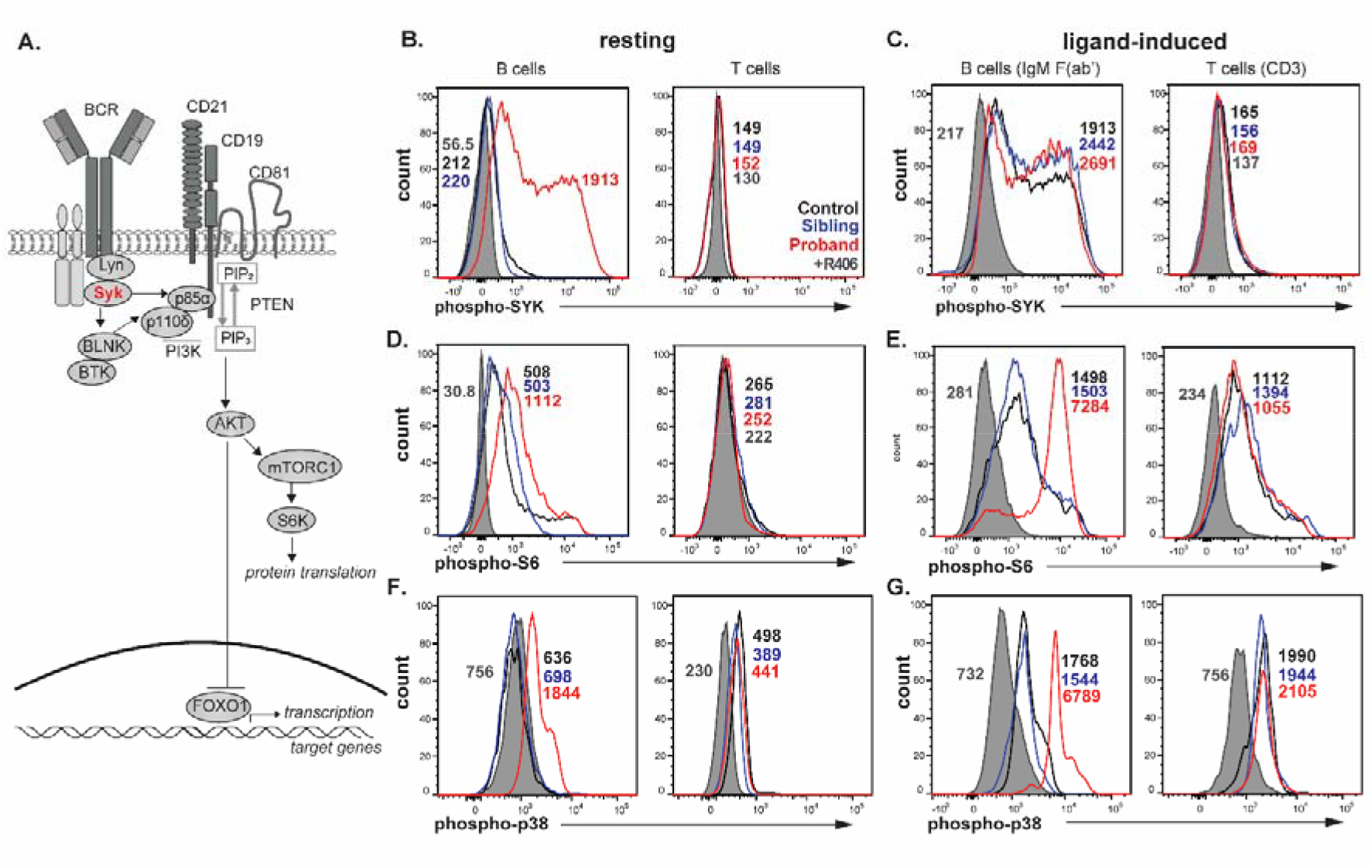
Functional impact of *SYK* variants on antigen-receptor signaling in B and T cells. (A) Schematic of the PI3K signaling pathway downstream of the B-cell receptor. (**B**) Resting and (**C**) ligand-induced levels of phospho-SYK (Tyr525/526) in B cells (anti-IgM F(ab’)_2_) and T cells (CD3). (**D**) Resting and (**E**) ligand-induced levels of phospho-S6 (S235/S236) in B and T cells. (**F**) Resting and (**G**) ligand-induced levels of phospho-p38 (T180/Y182) in B and T cells. Shown are the proband (*red*), unaffected sibling (*blue*) and a healthy control (*black*), and the proband cells treated with SYK specific-inhibitor R406 (*shaded grey*). Numbers on plots represent the median fluorescence intensity (MFI) levels.

The functional consequence of increased SYK autophosphorylation on downstream PI3K and MAPK signaling was examined through detection of phospho-S6 and phospho-p38 (**Figure 2D-G)**. Resting B cells from the patient showed 2-fold higher levels of phospho-S6 and 2.5-fold higher phospho-p38 than both the healthy control and unaffected sibling, whereas both were undetectable in resting T cells of all individuals (**Figure 2D&F**). Upon BCR stimulation, phospho-S6 and phospho-p38 levels were increased in B cells of all individuals, with those of the patient being 4.8-fold and 3.8-4.4-fold higher, respectively (**Figure 2E&G**). Phospho-S6 and phospho-p38 levels in T cells were similarly increased upon CD3 stimulation in all individuals (**Figure 2E&G**). B- and T-cell stimulation with PMA, which bypasses SYK, induced similarly high phospho-S6 levels in the patient, the unaffected sibling and healthy control (**Figure E1**). This demonstrates that the observed functional effects on both phospho-SYK and S6 are directly attributable to the identified *SYK* variant in this patient. Patient cells were also treated with PI3K inhibitor LY294002 to examine its capacity to dampen enhanced phosphorylation SYK, S6 and p38 (**Figure 3**). Whilst treatment with R406 effectively normalizing signals for phospho-SYK, phospho-S6 and phospho-p38, LY294003 only abrogated phosphorylation of S6 but not phospho-SYK and phospho-p38. Thus, the p.R590Q variant causes SYK autophosphorylation at Tyr525/536, which drives enhanced downstream signaling as evidenced by increased basal phosphorylation of S6 and p38 (**Figure 2**). These abnormalities can be effectively normalized by SYK inhibitor R406, but not PI3K inhibitor LY294002.

**Figure 3:**
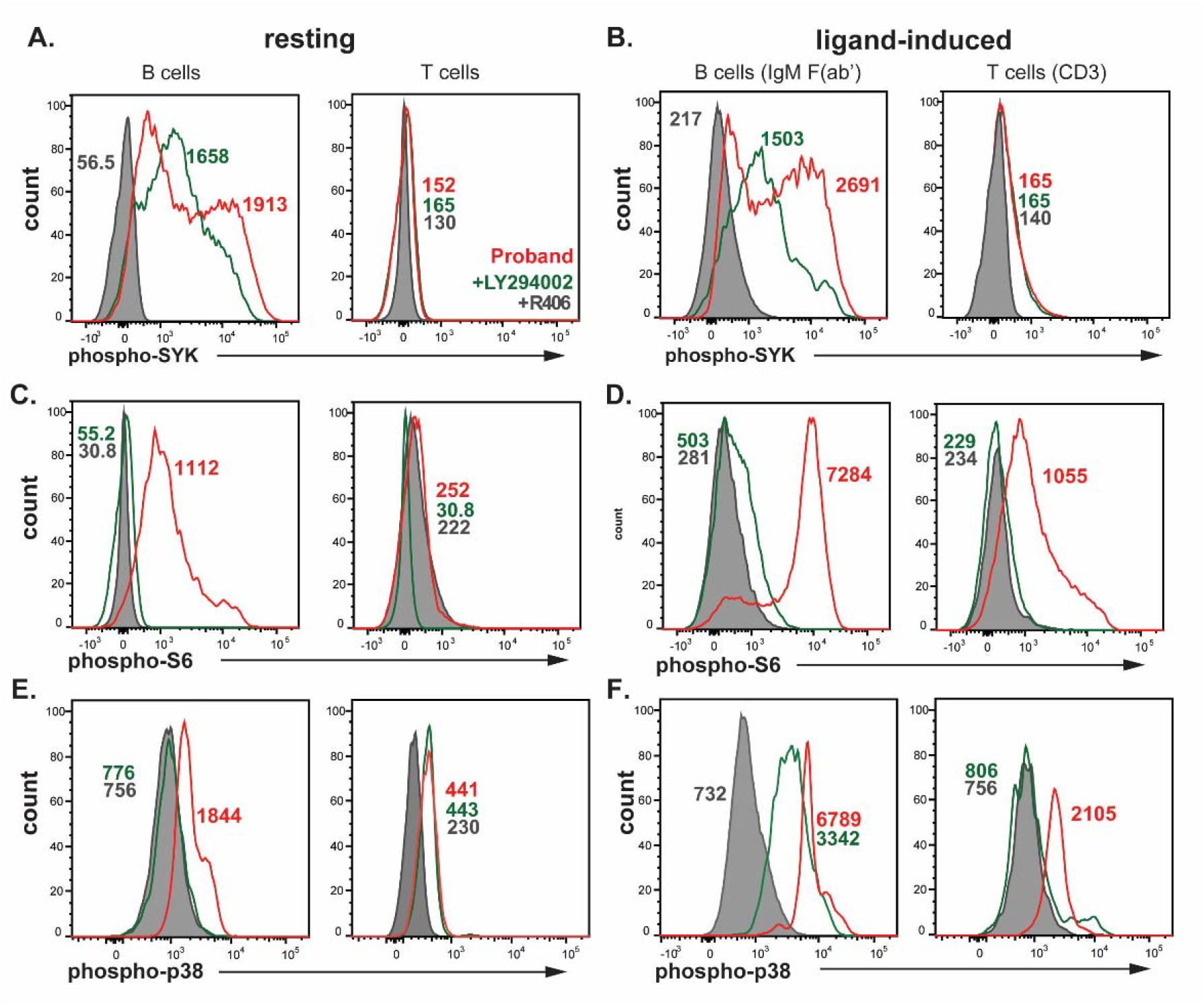
Impact of PI3K inhibition on enhanced antigen-receptor signaling. (**A**) Resting and (B) ligand-induced levels of phospho-SYK (Tyr525/526) in B cells (anti-IgM F(ab’)_2_) and T cells (CD3). (**C**) Resting and (**D**) ligand-induced levels of phospho-S6 (S235/S236) in B and T cells). (**E**) Resting and (**F**) ligand-induced levels of phospho-p38 (T180/Y182) in B and T cells. Untreated patient cells are shown in *red*, PI3K specific-inhibitor LY294002-treated cells (*green*) and SYK inhibitor R406 (*shaded grey*). Numbers on plots represent the median fluorescence intensity (MFI) levels.

**Figure 4:**
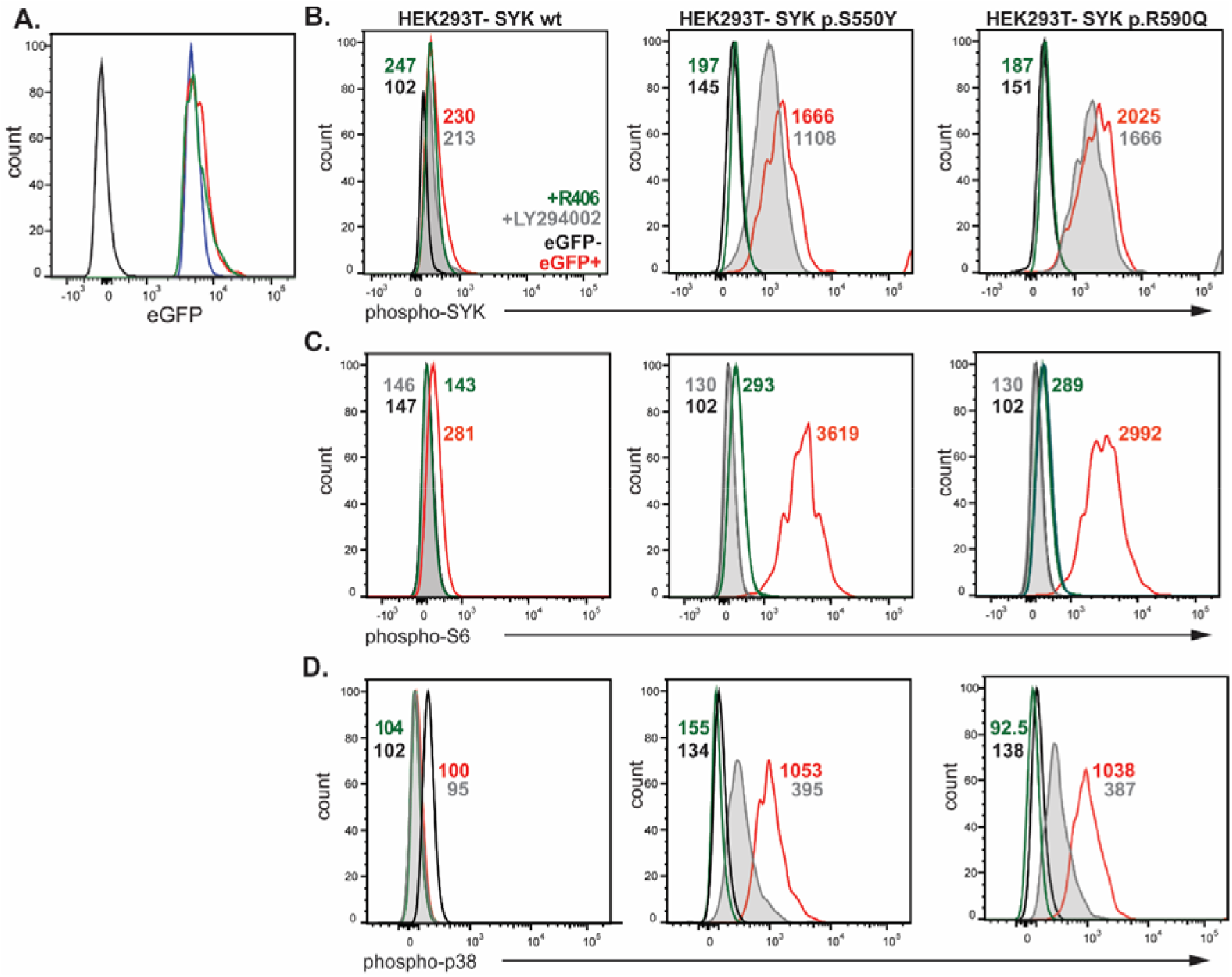
Functional validation of *SYK* variant in HEK293T cells transfected with wt or mutant SYK (p.S550Y[12] and p.R590Q) constructs. **(A)** eGFP expression of transfected HEK293T cells; untransfected (*black*), HEK293T-SYK wt (*blue*), HEK293T-SYK p.S550Y (*green*) and HEK293T-SYK p.R590Q (*red*). Levels of **(B)** phospho-SYK (Tyr525/526), **(C)** phospho-S6 (S235/S236) levels and (**D**) phospho-p38 (T180/Y182) following transfection in unstimulated HEK293T cells. Transfected cells were treated with PI3K specific-inhibitor LY294002 (shaded grey) or SYK inhibitor (green). Numbers in plots represent median fluorescence intensity (MFI) levels.

Causality between the novel *SYK* variant and the SYK autophosphorylation and PI3K activation was evaluated in the HEK293T cell line, which does not endogenously express SYK.[12] Expression constructs containing either wild-type (WT) SYK, the p.R590Q variant or the published p.S550Y variant, alongside an internal ribosome entry site-enhanced green fluorescent protein (IRES-eGFP) were introduced by transfection. All HEK293T-transfected cells displayed an equivalent median fluorescence intensity of eGFP, implying equal transcription levels (**Figure 3A**). While cells transfected with wild-type SYK showed low levels of phospho-SYK, phospho-S6 and phospho-p38, these levels were increased following transfection with the variant constructs (**Figure 3B,C**). Our results recapitulate those for p.S550Y,[12] and show similar functional consequences for the novel p.R590Q variant, as well as providing the first evidence that SYK GOF variants drive constitutive hyperactivation of the PI3K pathway. Together, these data demonstrate functional pathogenicity of *SYK* p.R590Q.

In summary, we present a patient with a novel missense variant in *SYK* causing constitutively high autophosphorylation of SYK, and activation of the PI3K and MAPK pathways, leading to hypogammaglobulinemia and multi-system inflammation. The observed autophosphorylation of SYK and hypogammaglobulinemia in our patient were reminiscent of observations from the six previously-described patients, as well as the *Syk*-Ser544Tyr mouse model.[12] Whilst all published SYK GOF cases exhibited multi-system inflammatory disease with the intestine and skin most recurrently affected, our case differed with inflammatory disease in the ovary, gut and lungs. Further, our patient displayed more severe lung complications with inflammatory pulmonary hypertension followed by severe fibrosis. Endothelin-1 mediated SYK-p38-MAPK pathway activation has been shown to modulate smooth muscle contraction [10, 11, 15], with SYK inhibition shown to attenuate muscle contractility [9]. As the SYK p.R590Q variant resulted in enhanced phospho-p38, we postulate that SYK GOF could increase airway endothelium contraction in the pulmonary vasculature contributing to development and/or severity of pulmonary arterial hypertension in our patient.[9]

We demonstrated for the first time that variants in SYK (both the novel p.R590Q and previously published p.S550Y)[12] result in constitutive hyperactivation and increased ligand-induced activity of the PI3K and MAPK pathways in B cells. The former is analogous to hyperactivation of the PI3K axis in patients with activated PI3K delta syndrome (APDS).[16, 17] As *in vitro* PI3K inhibition antagonized phospho-S6 but not phospho-SYK or phospho-p38 in our patient’s cells, treatment with pharmacological agents targeting PI3K, e.g. rapamycin or leniolisib, are unlikely to provide sufficient therapeutic benefit.[18, 19] SYK inhibition would be the preferred treatment strategy if available via off-label access, posing lower risks than a hematopoietic stem cell transplantation in adulthood.

By demonstrating the functional consequences of a novel variant in SYK on autophosphorylation of SYK, and PI3K and MAPK signaling we expand the genetic spectrum of SYK GOF. Furthermore, we demonstrate the capacity of functional evaluation of PI3K and MAPK signaling to enable validation of genetic variants for IEI diagnosis. As such assays could also guide patient stratification for targeted treatment, and be applied for monitoring of treatment efficacy.[20] Therefore, we advocate for embedding of this assay in healthcare supported diagnostics for IEI.

In summary, we present a patient with a monoallelic variant in SYK (p.R590Q) causing overactive PI3K and MAPK pathway function, hypogammaglobulinemia, some inflammatory disease and pulmonary arterial hypertension, with no evidence of recurrent infections.

For detailed methods, see Methods section of this article’s Online Repository.

## Data Availability

All data produced in the present study are available upon reasonable request to the authors

## Abbreviations

APDS: activated p110*δ* syndrome
BCR: B-cell antigen receptor
BLNK: B-cell linker protein
IBD: inflammatory bowel disease
IEI: inborn errors of immunity
PAD: predominantly antibody deficiency
PI3K: phosphoinositol-3-kinase
phospho-S6: phosphorylated ribosomal S6
SYK: spleen tyrosine kinase
LTx: lung transplantation
WES: whole exome sequencing.

## ACKNOWLEDGEMENTS

We are grateful to the patient for participating in this study and for providing approval to publish this case. We thank Dr Samar Ojaimi for her important discussions relating to this case, Ms Pei Mun Aui, Ms Fiona Hore-Lacy and Ms Ebony Blight for technical support. We acknowledge the services of the 3billion Inc, ARAFlowCore and Micromon facilities at Monash University.

## AUTHORS CONTRIBUTIONS

ESJE and MCvZ designed the study and wrote the manuscript. JC, GS and JJB provided patient care and clinical data. PMA, GHS and RK performed experiments. REO’H contributed to essential discussion and editing of the manuscript. All authors critically read and commented on the manuscript drafts and approved of the final version.

## Data availability

Data available on request

