## Supplemental material for "Novel *SYK* variant causes enhanced SYK autophosphorylation and PI3K activation in an antibody-deficient patient"

ONLINE REPOSITORY

**MATERIALS AND METHODS**
**Ethics**

Diagnostic examination of the patient included consent for collection of medical information, donation of a peripheral blood sample, and whole-exome sequencing. The study was carried out as part of a multi-center study (projects Alfred Health Ethics Committee, 198/18 and Monash University CF15/771-2015-0344). In addition, the patient and her sibling consented to publication of this report. Healthy control data were collected after written consent was obtained under approval of the Monash University Human Ethics Committee (study 2016-0289). All studies were conducted in accordance with the Declaration of Helsinki.

**Detailed clinical description of the patient**

A Caucasian female in her fourties born to non-consanguineous parents was referred for assessment of hypogammaglobulinemia following an incidental finding during assessment for lung transplantation (LTx) in the setting of severe pulmonary arterial hypertension. Background medical history was consistent with pulmonary arterial hypertension in the setting of delayed patent ductus arteriosus closure in her teens. Prior infection history was minimal with infrequent sinusitis and otitis, genital herpes, but no severe infections. There was no history of cutaneous disease or inflammatory bowel disease, nor a relevant family history. Medications at time of referral and assessment included desvenlafaxinemacitentan, metoclopramide, nizatidine, pantoprazole, sildenafil, warfarin and epoprostenol infusion.

Clinical examination noted digital clubbing, low set ears, clinical features of severe pulmonary hypertension, and basal lung crepitations. Initial laboratory investigations showed a normocytic anaemia with a hemoglobin of 106g/L, lymphocyte count of 1.13x10^9^/L, serum IgA 1.4 g/L, IgM 1.8 g/L and IgG 4.4 g/L. Serum protein electrophoresis did not reveal a paraprotein. Baseline CD3, CD4, CD8, CD19 and CD56 counts were normal. B-cell analysis showed decreased naive B-cell frequencies, with a corresponding increase in IgM-memory B cells. Switched memory B cells accounted for 15.8% of total B cells. Hepatitis B, C, HIV and CMV serology were all negative. Karyotyping did not detect any abnormalities. Chest CT did not reveal any signs of thymic hypoplasia or bronchiectasis, but did note subtle areas of diffuse ground glass opacity. Diagnostic vaccination studies with polysaccharide Pneumococcal vaccine (Pneumovax-23) revealed poor antibody responses.

The patient was recruited for further immunophenotyping and genomics, which subsequently revealed a *SYK* variant in exon 12, mapping to the protein kinase domain of the SYK protein (**Figure 1D**; see main text for more information).

Critically ill and bridged on extracorporeal membrane oxygenation, the patient subsequently proceeded to bilateral LTx. The surgery was complicated by massive hemorrhage, acute renal failure requiring hemofiltration, bilateral pulmonary embolism and gastroparesis. The patient recovered and post-transplant immunosuppression included tacrolimus, mycophenolate and prednisolone. Immunoglobulin replacement therapy was commenced post-transplant as infection prophylaxis in the setting of the hypogammaglobulinemia.

The native lung explant noted severe pulmonary hypertension changes with additional thromboembolism and unusual diffuse alveolar damage with hemorrhage. The patient subsequently returned to full health, although premature ovarian insufficiency, a colonic ulcer and pancreatic pseudocysts were noted. Three years post-LTx, a severe picornavirus pneumonia occurred, followed by progressive diffuse fibrotic lung disease and respiratory failure. She underwent retransplantion with a single right lung and doing well in the ~8 months follow-up.

**Flowcytometric Immunophenotyping**

Healthy control immunophenotyping data (n=59) were obtained over a 6-year period. Standardized sample preparation, antibody staining and flow cytometer instrument settings were used to ensure consistency of flow cytometric data.[1] Briefly, absolute counts of CD19^+^ B cells, CD3^+^, CD4^+^ and CD8^+^ T cells, and CD16^+^CD56^+^ NK cells were determined in whole blood using BD Trucount tubes (BD Biosciences, Franklin Lakes, NJ) and a lyse-fix protocol using BD FACS Lysing Solution (BD Biosciences) within 24h of blood sampling in EDTA-containing Vacutainers (BD bioscience). For detailed B- and T-cell immunophenotyping, peripheral blood was lysed in 0.155M NH_4_Cl for 15 minutes on a roller at room temperature, followed by centrifugation and washing twice with phosphate-buffered saline containing 0.5% bovine serum albumin and 10% sodium azide. As previously published, 1-2 million nucleated cells were incubated with antibodies at room temperature for 20 minutes, before washing twice and resuspending in FACS buffer.[1] Samples were acquired on the LSRII, LSRFortessa or LSR Fortessa X20 (BD Biosciences, San Jose, Calif). All data were acquired using FACS DIVA v8.0.1 (BD Biosciences) and analysed on FlowJo v10 software (BD Biosciences).

For analysis of phospho-SYK and phospho-S6, wells of a 96-well flat-bottomed plate were preincubated overnight at 4^o^C with 50µl of 1µg/ml CD3 (clone: OKT3, ThermoFisher Scientific) in sterile PBS. After incubation, wells were washed three times with sterile PBS. Freshly thawed PBMC were pre-incubated for 30 minutes with or without selective PI3K inhibitor, LY294002 at final concentration 500µM [2] or 2 SYK inhibitor (R406) [3] (Selleck Chemical, Houston,Tx) in RPMI 1640 media (Sigma Aldrich) containing 10% fetal calf serum (ThermoFisher Scientific) and 100 units/ml penicillin and 100 µg/ml streptomycin (Sigma Aldrich). Cells were then seeded at 1x10^6^ cells per well in a flat bottomed 96 well plate. Cells were either incubated with media only (Fluorescence Minus Once and media only controls), 5µg/ml CD28 (clone: CD28.2, Thermofisher Scientific) for wells precoated with CD3, 1 µg/ml IgM (Fc5µ) F(ab’)2 (Merck) or 1µg/ml PMA (Sigma Aldrich). For cells pretreated with LY294002 or R406, additional media containing the inhibitor was added to ensure cells were maintained in a final concentration of 500µM for the entirety of the stimulation process (2.5 hours, 37^o^C, 5% CO_2_). Cells were then spun at 4^o^C, and stained at 4^o^C for 30 minutes with a cell surface antibody cocktail containing CD3 BV711 (BD Biosciences), CD4 PC7 (Beckman Coulter), CD20 BV605, CD45RO FITC (both from Biolegend), CD8 BV510, CD45 PerCP Cy5.5, CD27 BV421, IgG PE, Ig A PE and fixable viability stain 700 (all from BD Biosciences) (Supplementary Table E3). Cells were washed twice with FACS buffer after cell surface staining. Cells were incubated at 4^o^C for 30 minutes with Cytofix/Cytoperm Buffer (BD Biosciences), then washed twice with FACs buffer and fixed at 4^o^C for 30 minutes with Phosphoflow Perm buffer III (BD Biosciences). Cells were then washed twice in perm/wash buffer (BD Biosciences), and subsequently stained at 4^o^C for 1 hour with either anti-phospho-SYK (Tyr525/526) PE (Cell Signaling Technologies, Danvers, MA), anti-phospho-p38 (T180/Y182) AF488 (BD) or anti-phospho-S6 (Ser 235-236) AF647 antibody (Cell Signaling Technologies). After incubation, cells were washed twice in BD Perm/Wash, cells were resuspended in FACs buffer and acquired using standardized compensation and quality settings [1] on a BD LSR II instrument. Data were acquired with FACS DIVA v8.0.1 software and analyzed using FlowJo v10 (both BD Biosciences).

**Whole Exome Analysis**

Genomic DNA was isolated from post-Ficoll granulocytes (GenElute Mammalian Genomic DNA Miniprep Kit, Sigma-Aldrich, St Louis, Mo). Samples were sent to 3billion Inc. (Seoul-si Gangnam-gu, South Korea), and subjected to WES. All exonic regions were captured by xGen Exome Research Panel v2 (Integrated DNA Technologies, Coralville, Iowa). Captured regions were sequenced with Novaseq 6000 (Illumina, San Diego, Calif). Raw data were aligned against the GRCh37/hg19 reference genome. Variant calling and annotation were undertaken using open-source tools and in-house software, including variant filtration, classification and variant scoring against the patient’s clinical phenotype.[4] Variants were prioritized according to the patient’s clinical phenotype and the American College of Medical Genetics (ACMG) guidelines.[5]

**PCR and Sanger sequencing of *SYK* exon 12**

Exon 12 of *SYK* was amplified from genomic DNA of the patient with newly-designed primers (forward, 5’-CCCAACTGACTCCAACATCACAGAT-3’ and reverse, 5’- AAGCGGGCACATTCCTGATTCAT-3’) and prepared for sequencing on an ABI PRISM 3130XL (Applied Biosystems, Foster City, Calif) in the Monash Micromon core facility.

**Cell lines and lentiviral reconstitution**

The HEK293T cell line, was cultured at 37^o^C in DMEM supplemented with 10% fetal bovine serum, 100 units/ml penicillin and 100 µg/ml streptomycin. HEK293T cells were seeded in petri dishes and cultured for 16-24 hours before transfection. One wildtype and two mutant SYK constructs were generated containing either of the patient-identified variants: pMIGR1-FLAG-SLP76-mut-p.S550Y and pMIGR1-FLAG-SLP76-mut-p.R590Q. All constructs were sequence-verified by Sanger sequencing. The SYK wt and mutant constructs were transfected into the HEK293T cell line (CRL-3216, ATCC) using calcium chloride precipitation.[6]

**Multispecies alignment of the SYK region containing identified variants**

Protein sequences for SYK from *Homo sapiens* (P43405), *Macaca mulatta* (F7GLV6), *Bos taurus* (A0A3Q1MHG9), *Ovis aries* (W5PD78), *Sus scrofa* (Q00655), *Equus Caballus* (A0A3Q2HPD6), *Oryctolagus cunisculus* (G1TDW9), *Mus musculus* (P48025) were obtained from Ensembl, and from *Rattus rattus* (Q64725) from NCBI. Multiple sequence alignment was performed using ClustalW (<https://www.ebi.ac.uk/Tools/msa/clustalw2/>).

**SUPPLEMENTAL TABLES** (n=3)

**Table E1: *SYK* variant properties and clinical presentation in our patient and previously-published cases**

|  | Wang et al., 2021 [3] | | | | | Our study |
| --- | --- | --- | --- | --- | --- | --- |
| Variant | p.P342T | p.A353T | p.M450I | p.S550F | p.S550Y | p.R590Y |
|  | c.1024C>A | c.1057G>A | c.1350G>A | c.1649C>T | c.1649C>A | c.1769G>A |
| Exon | 8 | 8 | 9 | 11 | 11 | 11 |
| dbSNP | rs1827861920 | rs200167353 | rs1304839707 | Rs1828636794 | Rs1828636794 | - |
| ClinVar | 989387 | 989236 | 989237 | 989386 | 989389 | - |
| ClinVar Clinical Significance | Likely pathogenic | Uncertain significance | Likely pathogenic | Conflicting interpretations of pathogenicity | Pathogenic | - |
| ClinGen Allele Registry | - | CA5119412 | CA373802363 | - | - | - |
| OMIM | 600085.003 | - | - | 600085.002 | 600085.001 | - |
| gnomAD allele frequency | N/D | 0.00002475 | 0.000007953 | N/D | N/D | N/D |
| Domain | Interdomain A | Interdomain A | Protein kinase | Protein kinase | Protein kinase | Protein kinase |
|  | De novo |  |  |  | De novo |  |
| CADD_PHRED | 25.5 | 27.3 | 26.3 | 29.7 | 28.3 | 25.8 |
| PolyPhen2 | Damaging | Damaging | Tolerated | Damaging | Damaging | Probably damaging |
| SIFT | damaging | tolerated | damaging | damaging | damaging | deleterious |
| No. patients identified to date | 1 | 1 | 1 | 2 | 1 | 1 |
| Basal pSYK (Tyr525/526) | increased | increased | increased | increased | increased | increased |
| Clinical Presentation |  |  |  |  |  |  |
| Age at diagnosis | 12yrs | 44yrs | 34yrs | 2wks | 2wks | 40-45yrs |
| Hypogammaglobulinaemia | + | + | + | + | + | + |
| Recurrent Infections | + | + | + | + | + | - |
| Intestinal inflammation | + | + | + | + | + | + |
| Skin inflammation | + | - | + | + | + | - |
| Joint inflammation | - | + | - | + | + | - |
| Lung inflammation | - | + | + | +/- | - | ++++ |
| Central nervous system inflammation | + | - | + | - | - | - |
| Liver inflammation | - | + | - | - | - | - |
| DLBCL | - | + | + | - | - | - |
| Pulmonary hypertension | - | - | - | - | - | + |
| N/D not detected; DLBCL, diffuse large B-cell lymphoma; CADD, Combined Annotation Dependent Depletion, gnomAD, Genome Aggregation Database; OMIM, Online Mendelian Inheritance in Man; SIFT, Sorting Tolerant from Intolerant; SYK, spleen tyrosine kinase.  CADD scores are a ranking whereby higher scores are more likely to be deleterious. Scores >20 are predicted to be amongst the top 1% most deleterious possible substitutions in the human genome.[7] | | | | | | |

**Table E2: Antibody list**

| **Marker** | **Fluorochrome** | **Clone** | **Source** | **Cat. Number** | **volume/**  **100μl test (in μl)** | **tube(s)** |
| --- | --- | --- | --- | --- | --- | --- |
| CD3 | FITC | UCHT1 | BD Biosciences | 555332 | 3 | 1 |
| CD3 | BV711 | UCHT1 | BD Biosciences | 563725 | 2.5 | 3,4,5,6,7 |
| CD4 | PC7 | SFCI12T4D11 | Beckman Coulter | 6607101 | 0.2 | 1,5,6,7 |
| CD4 | BV510 | RPA-T4 | Biolegend | 300546 | 1.5 | 3,4 |
| CD5 | APC | UCHT2 | Biolegend | 300612 | 0.2 | 2 |
| CD8A | APC-H7 | SK1 | BD Biosciences | 560179 | 4 | 1,3,4 |
| CD8A | BV510 | SK1 | BD Biosciences | 563919 | 2.5 | 5,6,7 |
| CD16 | PE | B73.1 | Biolegend | 360704 | 0.2 | 1 |
| CD19 | APC | SJ25C1 | Biolegend | 363006 | 0.4 | 1 |
| CD19 | PE-CY7 | SJ25C1 | BD Biosciences | 557835 | 5 | 2 |
| CD20 | BV605 | 2H7 | BD Biosciences | 563783 | 2.5 | 5,6,7 |
| CD21 | BV711 | B-ly4 | BD Biosciences | 563163 | 5 | 2 |
| CD24 | APC-Alexa750 | ALB9 | Beckman Coulter | B10738 | 2.5 | 2 |
| CD25 | BV421 | BC96 | Biolegend | 302630 | 2.5 | 4 |
| CD27 | BV421 | M-T271 | BD Biosciences | 562513 | 1 | 2,3,5,6,7 |
| CD28 | PerCP-Cy5.5 | CD28.2 | Biolegend | 302922 | 5 | 3 |
| CD31 | PE | WM59 | BD Biosciences | 555446 | 5 | 3 |
| CD38 | BV605 | HB7 | BD Biosciences | 562665 | 0.2 | 2 |
| CD45 | PerCP-Cy5.5 | 2D1 | BD Biosciences | 340953 | 2 | 1 |
| CD45RA | BV605 | HI100 | Biolegend | 304134 | 0.2 | 3,4 |
| CD45RO | FITC | UCHL1 | Biolegend | 304204 | 5 | 3,5,6 |
| CD56 | PE | B159 | BD Biosciences | 555516 | 5 | 1 |
| CD127 | APC | A019D5 | Biolegend | 351316 | 5 | 4 |
| CCR4 | PE-CY7 | L291H4 | Biolegend | 359410 | 5 | 4 |
| CCR6 | PerCP-CY5.5 | G034E3 | Biolegend | 353406 | 2.5 | 4 |
| CCR7 | PE-Dazzle | G043H7 | Biolegend | 353236 | 2.5 | 3,4 |
| *CCR7 | PE-CF594 | 150503 | BD Biosciences | 562381 | 5 | 3,4 |
| CXCR3 | PE | 1C6/CXCR3 | BD Biosciences | 557185 | 20 | 4 |
| CXCR5 | BB515 | RF8B2 | BD Biosciences | 564624 | 5 | 4 |
| HLA-DR | APC | L243 | Biolegend | 307610 | 2 | 3 |
| IgA | VioBright FITC | IS11-8E10 | Miltenyi Biotec | 130-104-726 | 0.5 | 2 |
| IgA | PE | IS11-8E10 | Miltenyi Biotec | 130-093-128 | 0.5 | 2,5,6,7 |
| IgD | PerCP-Cy5.5 | IA6-2 | Biolegend | 348208 | 1.5 | 2 |
| IgE | FITC | goat polyclonal | Invitrogen | H15701 | 1 | 2 |
| IgG | PE | G18-145 | BD Biosciences | 555787 | 5 | 2,5,6,7 |
| IgM | BV510 | MHM-88 | Biolegend | 314522 | 1 | 2 |
| pP38  (T180,Y182) | AF488 | p36/p38 (pT180/pY182) | BD | 612594 | 10 | 7 |
| pS6 (Ser235/S236) | APC | S535-536 | Cell Signalling | 14733S | 5 | 6 |
| pSYK (Tyr525/526) | PE | C87C1 | Cell Signalling | 6485S | 1 | 5 |
| TCRγδ | PC7 | IMMU510 | Beckman Coulter | B10247 | 1 | 3 |
| fixable viability stain 700 | AF700 | N/A | BD Biosciences | 564997 | 0.1 | 5,6 |
| * alternative reagent | | | | | | |

**Table E3: Composition of flow cytometry panels**

|  | **fluorochrome** | | | | | | | | | | |  |
| --- | --- | --- | --- | --- | --- | --- | --- | --- | --- | --- | --- | --- |
| tube | **BV421** | **BV510** | **BV605** | **BV711** | **FITC/ BB515** | **PerCP-Cy5.5** | **PE** | **PE-Dazzle** | **PC7 /  PE-Cy7** | **APC** | **AF700** | **APC-H7 / APC-Ax750** |
| 1. TruCount | **-** | **-** | **-** | **-** | CD3 | CD45 | CD16 + CD56 | **-** | CD4 | CD19 | - | CD8A |
| 2. B-cell | CD27 | IgM | CD38 | CD21 | IgE + IgA | IgD | IgG + IgA | **-** | CD19 | CD5 | - | CD24 |
| 3. T-effector | CD27 | CD4 | CD45RA | CD3 | CD45RO | CD28 | CD31 | CCR7 | TCRγδ | HLA-DR | - | CD8A |
| 4. Th subset | CD25 | CD4 | CD45RA | CD3 | CXCR5 | CCR6 | CXCR3 | CCR7 | CCR4 | CD127 | - | CD8A |
| 5. pSYK | CD27 | CD8A | CD20 | CD3 | CD45RO | IgD | pSYK | - | CD4 | - | Viability | - |
| 6. pS6 | CD27 | CD8A | CD20 | CD3 | CD45RO | - | IgG + IgA | - | CD4 | pS6 | Viability | - |
| 7. pP38 | CD27 | CD8A | CD20 | CD3 | pP38 | - | IgG + IgA | - | CD4 | - | Viability | - |

**SUPPLEMENTAL FIGURE**

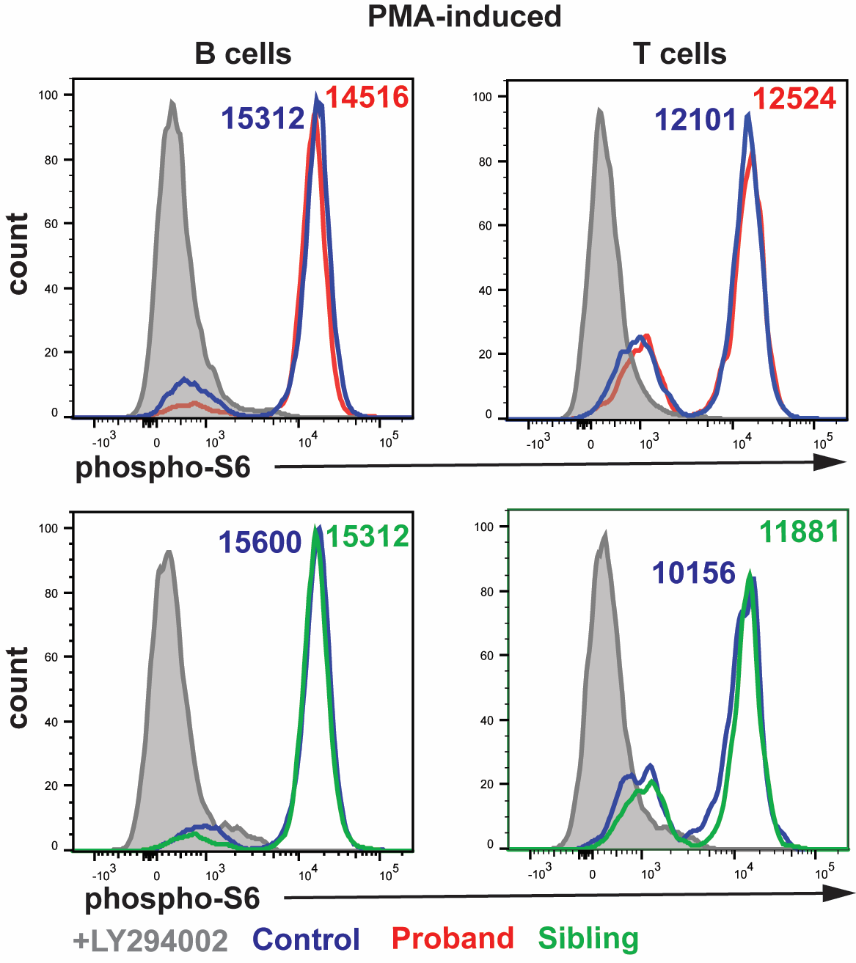

**Figure E1: PMA-induced PI3K signaling is intact in patient’s cells.** PMA induced levels of phospho-S6 levels in B cells and T cells of the affected patient (red), unaffected sibling (green) and a healthy control (blue). Shaded grey histogram represents patient cells treated with PI3K specific-inhibitor LY294002.
